# Camera-Agnostic Autonomous Diagnosis of Glaucomatous Optic Neuropathy using Macular Fundus Imaging and Machine Learning

**DOI:** 10.64898/2026.01.20.26344470

**Authors:** Zack Dvey-Aharon, Catherine Lalman, Tsontcho Ianchulev, Moshe Livne, Dan Margalit, Rachelle Aviv, Kristen Ann V. Mendoza, Joel S. Schuman

**Affiliations:** AEYE Health Inc., New York, NY, USA; Wills Eye Hospital, Philadelphia, PA, USA; Sidney Kimmel Medical College at Thomas Jefferson University, Philadelphia, PA, USA; New York Eye and Ear of Mount Sinai, Icahn School of Medicine, New York, NY, USA; Drexel University School of Biomedical Engineering, Science and Health Studies, Philadelphia, PA, USA; Vickie and Jack Farber Vision Research Center, Wills Eye Hospital, Philadelphia, PA, USA

**Keywords:** Glaucoma, fundus photography, camera-agnostic imaging, artificial intelligence, validation study

## Abstract

**Purpose:** Glaucoma, a leading cause of irreversible vision loss, often remains undiagnosed due to its asymptomatic progression and the limitations of existing screening methods. This study aimed to validate an artificial intelligence machine learning algorithm for the camera-agnostic detection of glaucomatous optic neuropathy using macula-centered fundus images.

**Methods:** Data were collected from EyePACS, a teleretinal screening system, comprising 25,000 macula-centered fundus images from 12,500 patients at U.S. primary care centers. A secondary dataset from the Philadelphia Telemedicine Glaucoma Follow-up Study was used for independent validation. A convolutional neural network was developed to detect glaucomatous optic neuropathy. Expert-graded fundus images served as the ground truth. Images underwent quality filtering to ensure the visibility of the optic nerve. Bilateral images were analyzed to produce patient-level diagnoses. Validation involved a secondary dataset of fundus images.

**Results:** The sensitivity and specificity of the algorithm in detecting glaucomatous optic neuropathy is calculated in comparison to expert grading. From the EyePACS dataset, 21,792 images (10,986 subjects) met quality standards. The algorithm demonstrated a sensitivity of 90.6% and specificity of 90.5%. Validation on the secondary dataset (200 fundus images from 100 subjects) resulted in a sensitivity of 96.4% and specificity of 85.3%.

**Conclusions:** The algorithm achieved high sensitivity and specificity in detecting glaucomatous optic neuropathy using macula-centered fundus images, demonstrating its potential for integration into diverse clinical settings. Its camera-agnostic design and robust performance offer a scalable solution for improving glaucoma screening pathways, making them more accessible and efficient.

## Introduction

Glaucoma is a leading cause of irreversible vision loss, affecting an estimated 90 million people worldwide. Characterized by progressive, permanent, and usually asymptomatic neurodegenerative loss of retinal ganglion cells that form the nerve fiber layer (RNFL) and optic nerve, half of all glaucoma cases remain undiagnosed^1^. Furthermore, many patients present late enough in the course of their disease that significant permanent optic nerve damage and visual field loss has already occurred^2–4^, making glaucoma the leading cause of irreversible blindness worldwide. Retinal injury can be slowed or prevented with timely treatment. Accordingly, accurate, efficient, and affordable glaucoma detection is essential. Unfortunately, glaucoma screening programs are currently limited by several complexities, including the relatively low prevalence of glaucoma and lack of cost effectiveness^5^. While a variety of modalities for glaucoma screening have been introduced, the most accessible and cost-efficient remains fundus photography. Here we report the results and predictive efficacy of an artificial intelligence (AI) trained algorithm for glaucomatous optic neuropathy (GON) based on camera-agnostic macula-centered fundus imaging using standard of care fundoscopy-based ground truth.

## Materials and Methods

A convolutional neural network (CNN) was trained to detect glaucoma from a single fundus image. An original model was trained using proprietary data with an Adam optimizer and a learning rate of 0.003. The model was trained on 2 GTX 2080 Ti graphic cards. The hyperparameters for the model training were chosen beforehand and not changed to prevent overfitting. After training, the CNN’s parameters were fixed, and the model was used to extract feature vectors from the right and left eye images of each patient. These two vectors were concatenated, meaning they were combined to create a single combined feature input. This input was passed into a simple fully connected neural network, commonly referred to as a perceptron, which made the final binary classification decision. As this study is a validation study of a pre-trained system. No optimization process was done. Additionally, single runs on the data were conducted per dataset. A schematic overview of the process is shown in Figure 1. The network consists of an initial “stem” convolutional block followed by a series of modular building blocks (Modules 1-5) that combine standard and depthwise convolutions, batch normalization, and nonlinear activations. These modules are arranged into sub-blocks with residual (“Add”) connections and repeated across seven main blocks to extract increasingly abstract features from macula-centered fundus images. Global average pooling and final convolutional layers produce a compact feature representation for each eye that is then used by the bilateral decision layer. Model architecture may be found in supplementary Figure 1.

**Figure 1.**
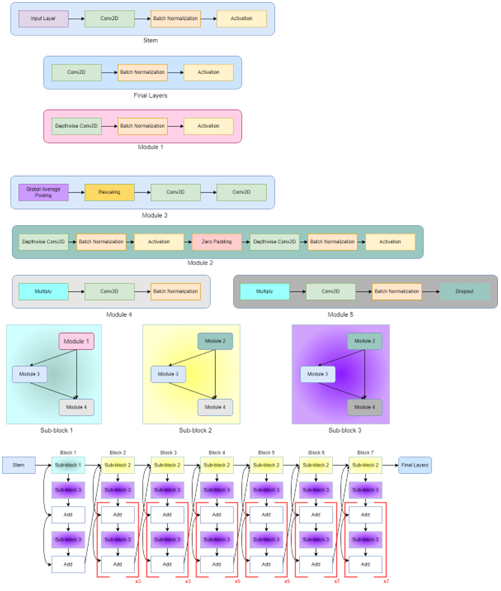
Schematic architecture of the convolutional neural network used in the AEYE glaucoma detection system.

Validation data were obtained from EyePACS, a teleretinal screening service which has constructed a database comprised of fundus retinal images and expert readings of said images. The data consisted of 25,000 images from 12,500 patients who visited one of the American primary care clinics using the EyePACS system; Table S1 summarizes the demographics of the cohorts. Although the ethnic distribution is not comparable to the population of the United States, where data were collected, the overall number of images provides useful data for all main US demographic groups. Six different cameras were used to obtain fundus photos: Topcon NW-400 (Topcon Healthcare, Oakland NJ, U.S), Canon CR-2 (Canon Medical Systems, Irvine CA, U.S.), CenterVue DRS (Optimetrics Inc., Alexandria VA, U.S.), CrystalVue (CrystalVue Medical Corporation, Taoyuan, Taiwan, China), Zeiss Visucam (Zeiss, Brochin, Germany), and Nidek AFC-330 (Nidek Inc., San Jose, CA, U.S.); acquisition protocols followed standard EyePACS procedures, as previously described^6, 7^.

EyePACS has a standardized internal process of expert consultation and validation for the diagnosis of GON^6^. Ophthalmologists first indicate and categorize cases according to the presence of referable glaucoma determined by analysis of fundus photos, where anatomical changes such as cup-to-disc ratio, asymmetry, notching, and disc hemorrhage were considered in order to diagnose referable GON. Each patient had 6 undilated fundus images and 2 anterior segment images taken, unless the pupil were small and imaging accordingly inadequate, in which case either one or both pupils were dilated and re-imaged^7^. There was no additional information available about the patients’ experience being imaged. The ground truth was then determined using 6 fundus images and 2 anterior segment images, where glaucoma was diagnosed by examining each pair of eye images. There was no additional characterization or quantification of severity of GON. Additional pathological characteristics are documented by graders and included in the analysis set as metadata. Because the EyePACS database was constructed primarily for screening purposes, the vast majority of participants had no prior documented eye disease diagnosis; cases with coexisting retinal disease or media opacity were retained and not excluded from this analysis in order to better approximate real-world screening conditions. These classifications were used as the ground truth in this research. Of the 12,500 patients analyzed, 2,250 were found to have referable glaucoma. It should be noted that the relatively high prevalence of glaucoma in this dataset, as well as the relatively low average age, may be explained by the fact that many patients were referred for screening or vision issues and by the fact that the original research dataset was enriched for higher glaucoma prevalence. These factors result in an effective age lower than the population average. Table S2 provides a summary of data sources and methodology.

For the present analysis, we included EyePACS participants who had a complete set of de-identified retinal photographs and expert gradings for glaucomatous optic neuropathy and whose images passed the automated quality filters described below. Patients were excluded if either eye lacked a gradable image or if required grading information was missing. No patients were excluded on the basis of ocular comorbidities or media opacities, in order to better approximate real-world screening conditions.

Prior to diagnosis performed by the AEYE algorithm, two types of image quality filters, a basic filter, which ensured that the optic nerve was visible in both images, and an additional filter which applied a further quality analysis score were applied to images centered on the optic nerve. No disease or media opacity were reasons to exclude from the study; exclusion was determined solely by the automated quality assessment of the images. Application of these filters in 11,702 (93.6%) and 10,896 (87.2%) patients whose images were deemed of sufficient quality, respectively. Qualified images were randomly selected for training purposes. These image sets were then uploaded directly to the cloud-based AEYE Glaucoma diagnostic screening system. For all analyses, the AEYE system produced a single patient-level classification based on the combined evaluation of both eyes. Images were split on the patient level, ensuring no patient appeared in more than one data subset (training or validation).

A second validation was performed on a dataset of 200 fundus photos taken during The Philadelphia Telemedicine Glaucoma Follow-up Study at Wills Eye Hospital, Philadelphia, PA, USA, using a nonmydriatic fundus camera (Volk Optical, Mentor, Ohio, USA)^8^. During the study, 906 individuals had 4 photos taken, two of which were fundus images that were centered on the optic nerve head, and two of which were centered on the macula. The study did not note if these images were dilated or undilated images, nor did it record any details regarding the patients’ experience while the photos were taken. Notably, of the 906 participants, 11.3% were also diagnosed with diabetic retinopathy, and 7.5% diagnosed with other retinal abnormalities^8^. A sample of 100 randomly chosen subjects, 46 of whom were healthy, and 54 of whom had signs of referable glaucoma, was studied. For this part of the analysis, fundus photos centered on the macula from these subjects were selected, and only subjects with a complete pair of gradable macula-centered images (one per eye) were included in the external validation cohort. Fundus photos centered on the macula from these subjects were analyzed by the algorithm. Demographics are shown in Table S1, and the process of cohort construction and quality filtering are shown in Figure 2.

**Figure 2.**
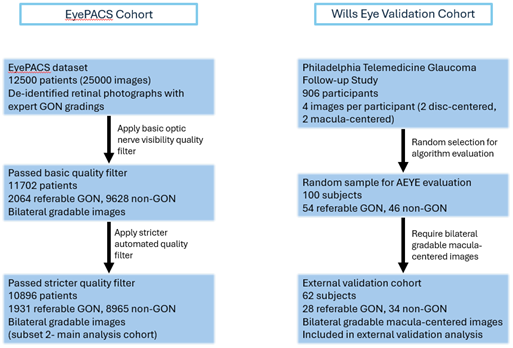
For the EyePACS dataset (left branch), 12,500 patients (25,000 images) were initially available. After application of a basic optic nerve visibility quality filter, 11,702 patients (2,064 with referable glaucomatous optic neuropathy [GON] and 9,638 without GON) had bilateral gradable images and comprised the first quality-filtered subset. Application of a stricter automated quality analysis filter further reduced the cohort to 10,896 patients (1,931 with referable GON and 8,965 without GON), which served as the main analysis cohort reported in the Results. For the Philadelphia Telemedicine Glaucoma Follow-up Study at Wills Eye Hospital (right branch), 906 participants were originally imaged. From these, 100 subjects (54 with referable GON and 46 without GON) were randomly selected for algorithm evaluation. Among them, 62 subjects (28 with referable GON and 34 without GON) had bilateral gradable macula-centered images and were included in the external validation analysis.

Although the Philadelphia study includes the term “telemedicine” in its title, the protocol did not involve remote or at-home screening. Instead, the study was conducted in physical community health settings, where trained personnel acquired fundus images that were later reviewed asynchronously. As such the validation cohort more accurately reflects a community-based outreach program incorporating elements of teleophthalmology through remote image interpretation.

Two images of each subject were analyzed at once – one from each eye – such that presence of GON^9^ was diagnosed per-patient rather than per eye, due to the importance of doing both-eye analysis in the diagnosis of GON using the AEYE algorithm. Images were uploaded without metadata in order to mask diagnoses such that analyses were performed independent of established ground truth. In the case where a subject had one healthy eye and one eye with glaucomatous changes, the glaucoma specialist classified the subject as positive for glaucoma because the presence of disease in even one eye was sufficient to necessitate further evaluation and follow-up. This in turn, was comparable to the way the algorithm identified glaucoma suspects. The system’s results were then statistically compared to the output results of the ground truth arm. In this study, the AEYE system was used as a fixed, commercially available algorithm; the study authors did not modify or retrain the underlying model.

Statistical analysis. Diagnostic performance was evaluated at the patient level. Sensitivity, specificity, positive predictive value (PPV), and negative predictive value (NPV) were calculated from 2×2 contingency tables. Exact 95% confidence intervals (CIs) for these proportions were obtained using Clopper–Pearson binomial intervals and are reported in Table 1. For the EyePACS dataset, we additionally estimated PPV and NPV at a representative U.S. glaucoma prevalence of 1.62% using standard formulas based on the observed sensitivity and specificity.

**Table 1.** Diagnostic performance of the AEYE Glaucoma system on the EyePACS and Wills Eye cohorts at the patient level. “Referable GON” indicates eyes graded as having glaucomatous optic neuropathy warranting referral by human readers. Sensitivity, specificity, positive predictive value (PPV), and negative predictive value (NPV) are reported as percentages with 95% confidence intervals (CIs) calculated using exact Clopper–Pearson binomial intervals. All metrics are computed at the patient level using the fixed decision threshold of the commercial AEYE Glaucoma system. For the EyePACS cohort, additional prevalence-adjusted PPV and NPV at a representative U.S. glaucoma prevalence of 1.62% are provided in the Results.

| Dataset | Subset | N patients | Referable GON n (%) | Sensitivity % (95% CI) | Specificity % (95 % CI) | PPV% (95% CI) | NPV% (95% CI) |
| --- | --- | --- | --- | --- | --- | --- | --- |
| EyePACS | Basic quality filter | 11702 | 2064 (17.6%) | 90.2 (95% CI 88.9-91.5) | 90.2 (95% CI 89.6-90.8) | 61.0 (95% CI 59.3-62.8) | 97.7 (95% CI 97.3-98.0) |
| EyePACS | Stricter quality filter | 10896 | 1931 (17.7%) | 90.6 (95% CI 89.2-91.8) | 90.5 (95% CI 89.9-91.1) | 67.2 (95% CI 65.4-69.1) | 97.8 (95% CI 97.5-98.1) |
| Wills Eye | External validation | 62 | 28 (45.2%) | 96.4 (95% CI 81.6-99.9) | 85.3 (95% CI 68.9-95.0) | 84.4 (95% CI 67.2-94.7) | 96.7 (95% CI 82.8-99.9) |

Ethics approval: Institutional Review Board exemption was obtained from the Sterling Independent Review Board under a category 4 exemption for the EyePACS dataset. For the Philadelphia Telemedicine Glaucoma Follow-up Study dataset, the study was declared exempt by the Wills Eye Hospital Institutional Review Board; informed consent was waived. No minors were included in the Philadelphia Telemedicine Glaucoma Follow-up Study dataset. The EyePACS dataset was accessed for research purposes on July 25, 2021 and the Philadelphia Telemedicine Glaucoma Follow-up Study dataset was accessed on September 13, 2023. All data were fully de-identified prior to analysis and none of the authors had access to information that could identify individual participants during or after data collection. Access to the underlying de-identified images remains governed by EyePACS and Wills Eye Hospital data-use agreements and institutional policies, as outlined in the Data Availability statement. This study adhered to the tenets of Declaration of Helsinki.

## Results

Of the first subset, consisting of patients whose images passed the optic nerve visibility filter, 2,064 subjects were diagnosed with referable GON by ophthalmologists. The model displayed 90.2% sensitivity and 90.2% specificity in the diagnosis of this subset. Of the 10,986 patients whose retinal images passed the second quality filter, 1,931 were diagnosed with referable GON by ophthalmologists. The model displayed 90.6% sensitivity and 90.5% specificity in the diagnosis of this subset. The results are displayed in Table 1.

As shown in Figure 3, each point represents the pair of sensitivity and specificity obtained at a different decision threshold on the model’s output score when evaluated at the patient level. Moving toward higher sensitivity is associated with a gradual decrease in specificity. The compromise between model sensitivity and specificity is far from linear. Although the sensitivity levels of 65%-70% translate to >97% specificity, balancing the sensitivity and specificity results in values between 91% and 92%, producing > 90% sensitivity and specificity simultaneously. Additionally, given the prevalence of glaucoma in U.S. adults is 1.62%^10^, the positive predictive value and negative predictive value can be calculated to be 13.6% and 99.8% respectively. When the second dataset’s 100 subjects, who were randomly selected from a US-specific population, were analyzed, the program determined that 28 glaucoma and 34 healthy pairs of images sufficient quality when evaluated by its optic nerve visibility filter. Of these, the algorithm was able to correctly identify glaucomatous features in 27 out of the 28 glaucoma cases. Similarly, the program was able to recognize 29 out of the 34 normal subjects as healthy. Accordingly, with this dataset the algorithm demonstrated a sensitivity of 96.4% and a specificity of 85.3%, corresponding to a PPV of 84.4% and an NPV of 96.7; 95% Cis for these estimates are shown in Table 1.

**Figure 3.**
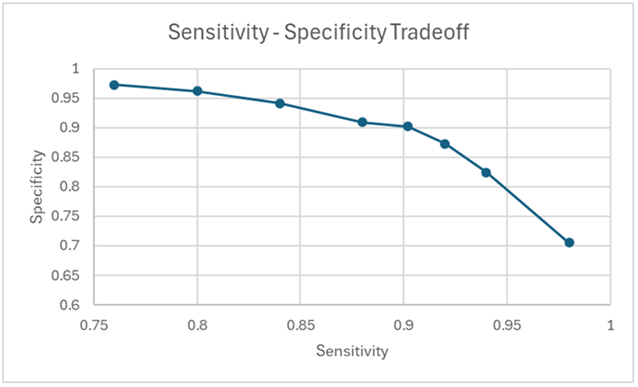
Sensitivity-specificity trade-off for the AEYE glaucoma detection algorithm.

## Discussion

Recently, deep learning algorithms, and convolutional neural networks in particular, have become increasingly accurate (compared to ground truth) in detecting glaucoma using fundus images. More specifically, machine learning algorithms have been developed to identify the presence of glaucoma by detecting small retinal structural changes. For example, one diagnostic hallmark of glaucoma is optic disc cupping, which represents its main clinical manifestation. Since cupping often precedes perimetric visual impairment, optic disc cupping and neuroretinal rim thinning have usually been the focus of screening and disease monitoring efforts.

Initial efforts for machine learning in glaucoma diagnosis by Ting et al. have demonstrated the feasibility of this approach^11^. Using a large database of 494,661 human-graded fundus photographs of diabetic patients, they used subjective grading by expert observers for referable GON, typically defined as optic disc abnormalities characteristic of glaucoma. Such abnormalities include but are not limited to: high cup:disc ratio; focally thin or absent neuroretinal rim, especially at the superior or inferior pole; and a vertically oval cup. The machine learning approach achieved sensitivity of 96.4% and specificity of 87.2% with an area under the receiver operating characteristic curve (AUC) of 0.942^12^. A later similar study by Li et al. reported an AUC, sensitivity, and specificity of 0.986, 95.6%, and 92%, respectively^13^.

In our study, the performance of our AI algorithm in the final validation dataset of 11,702 patients exceeds 90% sensitivity and specificity, comparable to those created by Liu et al and Ting et al.^11, 14^ The use of macula-centered images has an advantage over the disc-centered analysis of previois studies, as macula-centered images are easier to obtain with non-mydriatic camera. More specifically, the procedure needed to obtain a macular image is simpler for both the administrator and the subject. Furthermore, macula-centered images can also be analyzed for the presence of other diseases. ^17–24^ As such, these results present further opportunities for a synergistic streamlined screening system for comorbid macular and optic disc pathologies, including diabetic retinopathy, glaucoma, and AMD. AEYE Health, creator of the algorithm used in this study, for fundus-based diabetic retinopathy detection, suggesting that similar macula-centered pipelines can be deployed in routine clinical settings.^25^

Narrowing the ground truth of GON diagnosis to cup-to-disc ratio, as done in other studies, is problematic; previous studies have found that while important, cup-to-disc ratio is not the only crucial measure15. The variable features of the optic disc, such as differences in disc area, variability of cup-to-disc ratios associated with glaucoma, and pattern of retinal vessels which may confound disc analysis, make obtaining accurate, consistent photos for AI use challenging^11,16^. Additionally, analysis of the macula takes advantage of its relatively large size as a structure and ease of centration by focusing on the fovea with patient fixation to produce accurate, reproducible, and high quality images.

While many prior AI models for glaucoma detection have relied on optic disc-centered fundus images, our approach demonstrates that macula-centered images, which are more readily available in diabetic retinopathy screening and can achieve comparable or superior diagnostic performance. In the EyePACS dataset, the algorithm reached a sensitivity of 90.62% and specificity of 90.52%. Furthermore on an independent secondary validation set, performance remained strong with 96.4% sensitivity and 85.3% specificity. These results are on par with or exceed those reported in previous disc-centered models, which typically achieve AUCs in the range of 0.88-0.94 with sensitivity and specificity often below 90%^13, 14^. Moreover, our algorithm performed robustly on both macula- and disc-centered images, underscoring its adaptability to real-world clinical workflows.

## Limitations

Our study used the reference standard of ophthalmologist gradings, which may misdiagnose some cases and may not be as precise as formal reading center grading. As well, though the EyePACS dataset is large, which allows for reasonable representation of multiple ethnic groups, overall, the EyePACS screening population is not comparable to the US population. Additionally, SD-OCT data was not used in either the training or the validation datasets; use of OCT as a reference standard may increase the objectivity and quantification of the algorithm training, helping to further refine it. While a second follow-up visit including visual field testing was part of the original Philadelphia Telemedicine study protocol, only a small number of patients from the sample used to validate the AI algorithm attended this visit. Among them, just three had fundus images and completed visual field exams. Of these, two were excluded due to poor image quality, and the remaining patient was identified as a glaucoma suspect by both the AEYE algorithm and confirmatory testing. Although limited, this finding supports the algorithm’s potential clinical utility.

However, definitive diagnosis of glaucomatous optic neuropathy does require functional testing and that future validation using datasets with full confirmatory testing is warranted. Accordingly, our findings should be interpreted as detection of referable glaucomatous optic neuropathy based on fundus photography, rather than definitive diagnosis of early glaucoma. The absence of systematic structural (OCT) and functional (perimetry) confirmation in both datasets limits our ability to precisely stage disease severity or to quantify performance specifically for pre-perimetric glaucoma. However, most patients undergoing glaucoma screening today are not imaged using OCT, as this modality is not routinely included in population-level screening^26^. Looking ahead, the goal is to enable effective glaucoma screening without reliance on OCT-similar to the approach already established for diabetic retinopathy using fundus photography and other accessible tools.

Additionally, because the algorithm considers fundus image factors in addition to cup-to-disc ratio and unilateral abnormality, it was trained using pairs of images. Finally, disease severity was not a factor that was considered by the graders or evaluated by the algorithm. This in turn makes comparing this algorithm to its precedents more complex.

This study demonstrates the potential utility of a camera-agnostic AI-ML algorithm in the detection of glaucomatous optic neuropathy using macula-centered fundus images. Because the AEYE system is designed to operate on paired images from both eyes, our analysis is restricted to subjects with bilateral gradable photographs. This is in part due to the fact that the EyePACS ground truth was determined on the patient level and not the eye level. As a result, our results may over estimate performance in settings where one eye cannot be imaged due to media opacity, prior surgery, or other factors, and future work is needed to quantify performance under unilateral imaging conditions. With that said, the number of patients with images of a single eye was relatively low, implying that there is not a strong use-case for single-eye diagnosis. Furthermore, if a patient’s eye cannot be consistently imaged, a ophthalmological examination is recommended, negating the need for this system.

The external Wills Eye validation cohort was intentionally enriched for referable glaucomatous optic neuropathy, which is substantially higher than the estimated prevalence of glaucoma in the general population. While this enrichment improves statistical power in a relatively small sample, it also inflates apparent screening performance when metrics such as PPV are calculated at the cohort’s observed prevalence. To address this, we report PPV at a representative population prevalence of 1.6%, yielding a value of 13.6%, and we now emphasize that this more conservative estimate better reflects expected performance in true screening settings. Prospective population-based studies will be required to precisely quantify PPV and NPV under real-world prevalence.

## Data Availability

The datasets analyzed in this study consist of de-identified retinal images and associated clinical labels from the EyePACS teleretinal screening program and the Philadelphia Telemedicine Glaucoma Follow-up Study at Wills Eye Hospital. The authors do not own these datasets and are not permitted to redistribute the raw images. De-identified EyePACS images and labels are available to qualified researchers through EyePACS under a data-use agreement and data governance procedures; requests for access can be directed to EyePACS. De-identified images and labels from the Philadelphia Telemedicine Glaucoma Follow-up Study may be made available upon reasonable request to the corresponding author and are subject to approval by the Wills Eye Hospital Institutional Review Board and execution of an appropriate data-use agreement.

All aggregated results necessary to interpret the findings, including performance metrics and stratified analyses, are provided in the main text and Supporting Information. The internal source code and trained weights of the commercial AEYE Glaucoma system are proprietary to AEYE Health, Inc. and cannot be shared by the authors; the evaluation procedures and statistical analysis methods used in this study are described in detail in the Methods and are sufficient to allow independent assessment of similar screening workflows on other datasets. Researchers interested in testing the system may contact AEYE Heath to request system access.

