## Supplementary material for "Camera-Agnostic Autonomous Diagnosis of Glaucomatous Optic Neuropathy using Macular Fundus Imaging and Machine Learning": Table S1, Table S2

**Supplemental Data**

S1. Model workflow. A convolutional neural network (CNN) was trained to detect glaucoma for a single image as input. Following that and freezing the network (closing the network to further training), the features of both right and left eye were fed into a perceptron to decide the class.

**S1 Table. Statistical characteristics of the EyePACS and Wills Eye**	**Hospital dataset demographics**

| **Criterion** | **Distribution**  **EyePACS** | **Distribution**  **Wills Eye Hospital** |
| --- | --- | --- |
| **Age**  <40  41-60  >60  **Gender**  Male  Female  Other  **Ethnicity**  Caucasian  Latin American  African Descent  Indian Subcontinent  Asian  Native American  Multi Racial  Unspecified / Decline to State / Other | 11.2%  45.7%  43.1%  39.6%  56.9%  3.5%  16.2%  31%  9.1%  13.8%  4.4%  0.5%  2%  23.1% | **N/A**  **N/A**  **N/A**  **N/A**  **N/A**  **N/A**  7%  11%  74%  0%  7%  0%  0%  1% |

**S2 Table. Summary of data sources and methodology**	303

| **Data Source** | **Validation Set** | **Image Quality Assessment** | **Arm #1 –**  **Ground Truth** | **Arm #2 –**  **AI Algorithm** |
| --- | --- | --- | --- | --- |
| EyePACS | 12,500 Patients  (25,000 macula centered images, 25,000 optic nerve centered images) | Basic (visibility of the optic nerve) and advanced filtering configurations | US Experienced and Certified Ophthalmologist | AEYE Glaucoma algorithm  (Referable Glaucoma indication) |
| Wills Eye Hospital | 100 Patients  (200 optic nerve head centered images) | Basic (visibility of the optic nerve) and advanced filtering configurations | US Experienced Ophthalmologist | AEYE Glaucoma algorithm  (Referable Glaucoma indication) |
